# Potential of facial biomarkers for Alzheimer’s disease and Obstructive sleep apnea in Down syndrome and general population

**DOI:** 10.1101/2024.11.03.24316427

**Authors:** L.M Echeverry-Quiceno, S. Llambrich, Á. Heredia-Lidón, S. Giménez, M. Rozalem-Aranha, P. Inampudi, Y. Heuzé, X. Sevillano, J. Fortea, N. Martínez-Abadías, Alzheimer’s Disease Neuroimaging Initiative

**Affiliations:** Departament de Biologia Evolutiva, Ecologia i Ciències Ambientals (BEECA), Facultat de Biologia, Universitat de Barcelona (UB), Spain; HER - Human-Environment Research Group, La Salle - Universitat Ramon Llull, Barcelona, Spain; Multidisciplinary Sleep Unit - Respiratory Department, Hospital de la Santa Creu i Sant Pau, Biomedical Research Institute Sant Pau, Institut de Recerca Sant Pau -, Universitat Autònoma de Barcelona, Barcelona, Spain; Memory Unit, Department of Neurology, Institut de Recerca Sant Pau - Hospital de la Santa Creu i Sant Pau, Universitat Autònoma de Barcelona, Barcelona, Spain. Centro de Investigación Biomédica en Red en Enfermedades Neurodegenerativas (CIBERNED), Madrid, Spain; Neuroradiology Section, Department of Radiology, Hospital de la Santa Creu i Sant Pau, Universitat Autònoma de Barcelona, Barcelona, Spain; Columbia University College of Dental Medicine, New York, United States; Univ. Bordeaux, CNRS, Ministère de la Culture, PACEA, UMR5199, Pessac, France

**Keywords:** Trisomy 21, Facial development, Ontogenetic trajectories, Sexual dimorphism, Dementia, Sleep and respiratory disorders, Facial dysmorphology, Geometric morphometrics

## Abstract

**Background:** Down syndrome (DS) is caused by trisomy 21, leading to increased risks of Alzheimer’s disease (AD) and Obstructive sleep apnea (OSA). Traditional diagnostic methods for AD and OSA, like cerebrospinal fluid analysis and polysomnography, are invasive and challenging for people with DS. Facial morphology has emerged as a non-invasive biomarker for these conditions. Therefore, we conducted a comprehensive 3D analysis of facial shape variation in DS and euploid control (EU) populations, assessing the effect of critical factors such as age, sex, and facial size, to investigate its association with AD and OSA.

**Methods:** Facial shape differences among groups were analyzed from the coordinates of 21 landmarks automatically registered on 3D facial models generated from head magnetic resonance images in a cross-sectional sample of 131 individuals with DS and 216 EU controls from 18 to 90 years old, including subjects diagnosed with AD and OSA. We used Procrustes ANOVA and MANOVA tests to quantify the amount of facial shape variation attributable to sex, age, and facial size, and quantified facial shape differences among diagnostic groups using geometric morphometrics.

**Results:** Besides facial shape differences between DS and EU individuals, our results detected significant interactions between diagnosis and sex, and between diagnosis and age, indicating sex-dependent differences and an altered pattern of facial shape change over adulthood in DS, with females presenting more severe alterations. Multivariate regression analyses showed that facial shape significantly correlated with the concentration ratio between amyloid beta peptide 1- 42 and amyloid beta peptide 1-40 (Aβ1-42/Aβ1-40) in cerebrospinal fluid, a common biomarker for AD diagnosis. In the DS population, facial shape differences between individuals with and without AD diagnosis did not achieve statistical significance after adjusting for age and facial size, but significant shape differences were detected in the EU population. Regarding OSA, facial shape significantly correlated with the apnea-hypopnea index, and individuals with DS and severe OSA presented a significantly different facial morphology in comparison to individuals with DS and no signs of OSA, suggesting that facial morphology could be associated with sleep respiratory disturbances.

**Conclusions:** Overall, our study underscores the potential of facial morphology as a diagnostic biomarker for the early detection and clinical management of AD and OSA.

## Background

Down syndrome (DS) (OMIM 190685) is a complex genetic disorder occurring in one out of 700– 1,000 live births that is caused by trisomy of chromosome 21 [1]. The overexpression of more than 200 triplicated genes leads to a dosage imbalance, disrupting signaling pathways that regulate the development of multiple systems, including the nervous, cardiac, immune, gastrointestinal, and skeletal systems [2]. Advances in research, early intervention, educational therapy, and improvements in medical care have facilitated the management of pathologies associated with DS that manifest early in life. As a result, the quality of life of patients with DS has significantly improved, with life expectancy increasing from 9 to 60 years [1,3,4]. However, this increased longevity has led to greater vulnerability to age-related comorbidities [1,5], such as Alzheimer’s disease (AD) [6] and Obstructive sleep apnea (OSA) [7]. These disorders further impact cognitive impairment and substantially reduce quality of life in people with DS.

Individuals diagnosed with DS present an ultrahigh risk of developing early-onset AD, [6] In DS, the triplication of the *Amyloid Precursor Protein* (*APP*) gene, located in Hsa21 [8], is considered a necessary and sufficient condition to the overproduction of β-amyloid peptides (Aβ) in the brain and the development of AD [8]. Furthermore, individuals with DS also present a higher prevalence of severe sleep disruptions (74.5%) and OSA (78%) [7,9]. Typical features associated with DS, such as midfacial flatness and mandibular hypoplasia [10,11], muscular hypotonia [10,11], adenotonsillar hypertrophy and macroglossia [12], are risk factors for respiratory problems in people with DS. Sleep disorders have in turn been associated with cognitive deterioration due to deficient rest and lack of oxygen [13], as well as with higher burden and deposition rates of Aβ [14,15] and tau proteins [16,17] due to a poor protein clearance, further contributing to an earlier onset of AD in people with DS [18].

Diagnosis of AD and OSA is challenging in the general population, but particularly in individuals with DS. Research on biomarkers based on the analysis of protein concentration such as Aβ, tau and neurofilament light (NFL) in the cerebrospinal Fluid (CSF) showed a high diagnostic potential for AD [19,20]. However, CSF extraction is invasive and poorly tolerated by people with DS. Concerning OSA detection, an overnight polysomnography (PSG) study is the established gold standard [21]. However, this procedure can be tedious, cause discomfort, and has been questioned as a diagnostic tool in the DS population [12]. Similar to CSF extraction, performing a PSG in people with DS presents additional challenges, since the presence of craniofacial dysmorphologies, obesity and restless behavior during testing can alter the PSG output, misdiagnosing the presence and severity of OSA [21].

In the search of complementary biomarkers that are reliable and non-invasive to diagnose the presence of AD and OSA, facial morphology has emerged as a potential biomarker [22,23,24,25,26,27]. Recent advancements in imaging, computational, and artificial intelligence technologies have developed faster and more reliable tools for diagnosing genetic, rare, neurodevelopmental and psychotic disorders [22,23,26,28]. These tools use high-throughput methods based on 2D and 3D image analysis to identify phenotypic biomarkers through comparisons of facial patterns between patients and healthy controls [22,23,24,25,26,27]. Assuming facial development is closely integrated with brain and upper airway development through common signaling pathways [29,30], previous research underscored that facial traits may indirectly reflect neurological and respiratory disorders in the general population.

Specifically for AD, previous studies have demonstrated that automated computational methods can non-invasively detect dementia using facial traits, with an accuracy above 80% in 2D frontal images [31], and 66% with 3D facial meshes [32]. Regarding OSA, previous research has been able to detect this respiratory condition using non-invasive facial biomarkers with 2D images based on cephalometric parameters [33,34,35,36], as well as with 3D meshes, revealing significant local differences concentrated in the nasal region in patients with OSA [37]. Although these studies showed promising results that reinforce the reliability of using facial biomarkers for detecting AD and OSA, they overlooked significant factors that influence facial shape variation such as sex, age, and facial size. Moreover, these methods have not been assessed in individuals diagnosed with genetic conditions who are at an elevated risk of developing AD and OSA, such as the DS population.

To fill these gaps and to further improve the use of facial biomarkers for early detection and prognosis of neurodevelopment and respiratory disorders, we conducted the first comprehensive quantitative 3D analysis of facial shape variation including euploid controls (EU) and individuals diagnosed with DS, AD and OSA. First, we assessed the effect of age, sex, facial size and genetic factors on facial shape to understand how these critical factors modulate the facial phenotype. Second, we explored facial adult ontogeny in healthy controls and tested whether this process is altered in DS individuals depending on the sex. This is crucial in the investigation of the diagnostic potential of facial biomarkers because the mechanisms that regulate the process of facial development, growth and ageing vary during the ontogenetic trajectory, among individuals, and between sexes [38,39,40].

To address these issues, we applied quantitative geometric morphometric analyses [41,42,43], which have been previously used to identify disease and sex-specific facial features associated with genetic and neurodevelopmental disorders. In an earlier study, we reported subtle but significant facial shape differences between healthy controls and patients diagnosed with schizophrenia and bipolar disorder, showing that these facial traits were significantly different between disorders and between male and female patients within each disorder [28]. These results underscore the sensitivity of facial biomarkers for disease diagnosis. In this study, we accounted for the key factors modulating facial shape to assess the potential of facial biomarkers for detecting sex-specific facial traits associated to AD and OSA in the general and the DS populations.

## Methods

### Sample composition

The sample analyzed in this cross-sectional study comprised 347 individuals with and without diagnosis of AD, including 216 euploid individuals (EU) (72 males, 144 females), and 131 individuals with Down syndrome (DS) (80 males, 51 females) (Table 1). For all individuals, a head Magnetic Resonance image (MRI) was available, along with information regarding dementia stage and diagnosis of Alzheimer’s disease (AD). Data on Obstructive sleep apnea (OSA) was also available for a subsample of individuals (Table 1).

**Table 1.**
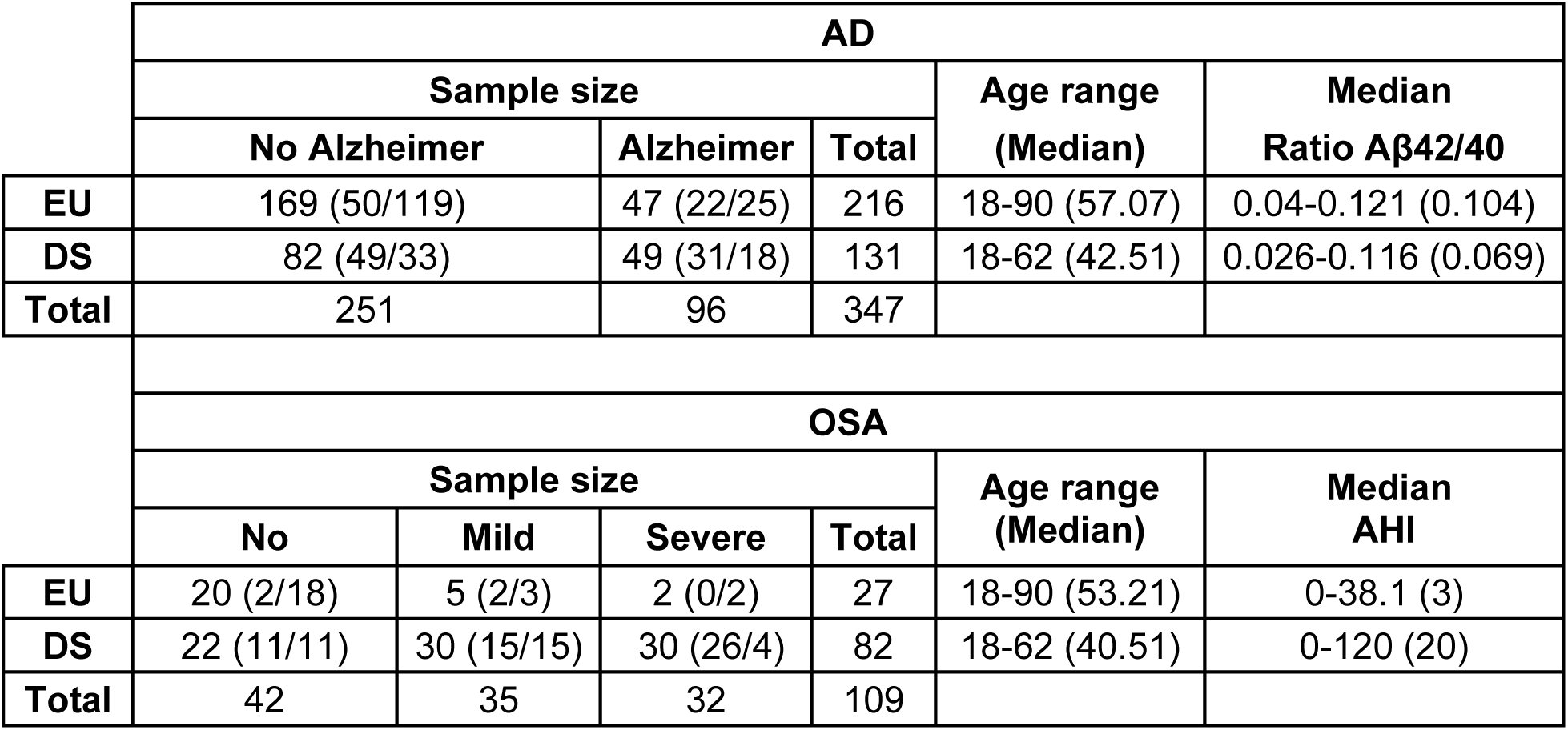
Sample composition of EU and DS groups divided by AD dementia stages and OSA level. The number of males and females is indicated in brackets as (Males/Females).

All individuals with DS were recruited from the population-based Down-Alzheimer Barcelona Neuroimaging Initiative (DABNI) cohort. Cognitively unimpaired EU participants were recruited from the Sant Pau Initiative on Neurodegeneration (SPIN) cohort, while EU participants with AD were selected from the Alzheimer’s Disease Neuroimaging Initiative (ADNI) database (adni.loni.usc.edu). All participants were of European origin, with ages ranging from 18 to 62 years (individuals with DS) and from 18 to 90 years (EU controls). The study was approved by the Sant Pau Ethics Committee following the standards for medical research in humans recommended by the Declaration of Helsinki. All participants or their legally authorized representatives gave written informed consent before enrollment. To use the ADNI database, we followed an application protocol that, after evaluation, provided us access to the clinical and imaging data of ADNI participants, who signed consent forms approved by the Resource Allocation Review Committee (RARC).

### AD diagnosis

In the DS cohort, AD diagnosis was established after neurological and neuropsychological examinations with a semi-structured health questionnaire (Cambridge Examination for Mental Disorders of Older People with Down Syndrome and others with intellectual disabilities [CAMDEX- DS]) [44], and a neuropsychological battery including the Cambridge Cognition Examination (CAMCOG). Both tests were adapted for intellectual disabilities, as in Fortea et al. (2018) [19]. In the ADNI EU cohort, AD diagnosis was obtained following the National Institute on Aging-Alzheimer’s Association guidelines for the neuropathologic and neuropsychological assessments of AD [45].

In both cohorts, the neuropsychological evaluations were accompanied by molecular data, including the concentration ratio of Aβ1-42/Aβ1-40 proteins obtained from CSF samples. For each individual, the degree of AD dementia was classified as “No Alzheimer” when no signs of AD dementia were detected, or as “Alzheimer disease” when individuals showed prodromal or advanced signs of AD dementia (Table 1), following the criteria in Fortea et al. (2018) [19].

### OSA diagnosis

To score the presence and severity of OSA, we used the apnea-hypopnea index (AHI) obtained from nocturnal polysomnography tests performed in sound-attenuated and temperature regulated rooms in the sleep laboratory at the Sant Pau sleep unit following the protocols described in Giménez et al (2018) [30]. AHI is defined as the sum of all apneas (airflow reduction greater than 90% for more than 10 seconds), and hypopneas (airflow reduction greater than 30% for at least 10 seconds with an oxygen saturation decrease of approximately 3% or a cortical awakening) per hour of sleep [7]. Individuals were classified into three categories according to the severity of sleep disruption: a) no OSA (AHI: <5), mild OSA (AHI: 5-30) and severe OSA (AHI: >30) (Table 1). To streamline the analysis and increase sample size for each group, we created a “mild OSA category” which contains two existing categories: mild OSA (AHI 5–15) and moderate OSA (AHI 15–30).

### Facial 3D models and landmarking

3D facial models were generated from MR images (Additional file 1: Figure. S1) by adjusting the grey-scale threshold that optimized skin segmentation using Amira 5.2.1 (v6.3, Visualization Sciences Group, FEI). The facial models showed similar reconstruction quality and were derived from T1-weighted images obtained from Hospital del Mar (Barcelona, Spain) or Hospital Clinic (Barcelona, Spain), and ADNI database. Since all individuals were scanned using a 3T scanner with similar scanning protocols (Additional file 2: Table S1), potential experimental bias due to different scan origin was considered negligible.

To capture facial shape, we used an automatic method to record the 3D coordinates of 21 anatomical landmarks on the 3D facial models generated from MR images (Fig. 1 and Additional file 3: Table S2). The automatic landmarking method is included in *BioFace3D*, a pipeline for the automatic computation of potential facial biomarkers from 3D head models reconstructed from MRI scans [46]. *Bioface3D* [46] applies a domain adaptation an architecture reuse approach [47] on Multi-View Consensus Convolutional Neural Networks (MV-CNN), a deep learning-based state-of-the-art method devised for automatic 3D facial landmarking [48]. This method is based on creating 2D views of the 3D anatomical facial models from different viewpoints. Then, the hourglass model is used to predict candidate landmarks on 2D heat maps, and the final position is determined by ray-projecting onto the surface of the 3D mesh using least squares combined with random sample consensus [48]. The model was trained with datasets of 21 landmarks manually collected on 538 facial meshes, including healthy controls and subjects with genetic and neurodevelopmental disorders [46]. This automatic facial landmarking method has an average landmarking error of 1.78mm. This accuracy level is lower than 2mm, which is the intra-observer measurement error commonly accepted for craniometric measurements [49].

**Fig. 1.**
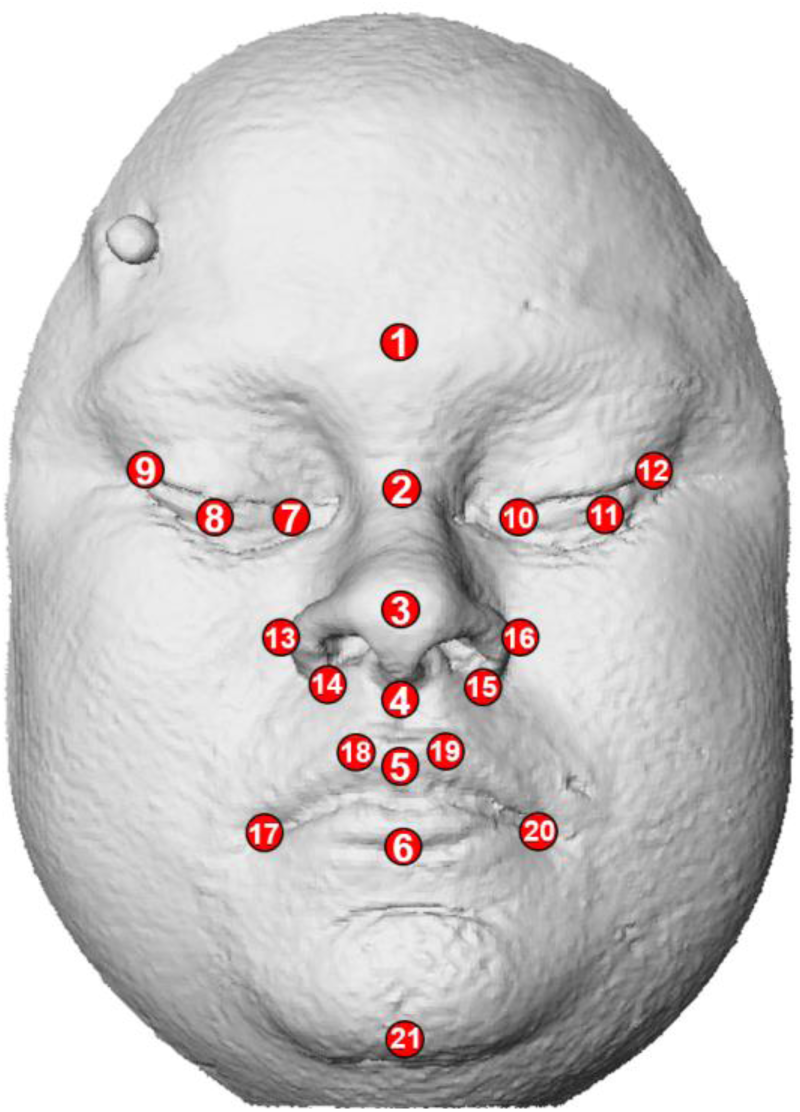
Anatomical position of the facial landmarks used for geometric morphometric and multivariate statistical analyses. For landmark definitions see Additional file 3: Table S2. This facial model is a morphing which does not belong to a real individual and is based on 3D landmark coordinates.

### Geometric Morphometrics and multivariate statistical analyses

To quantify facial shape variation, we used Geometric Morphometric (GM) analyses. First, we performed a Generalized Procrustes Analysis (GPA) [50] to superimpose all the configurations of landmarks into a common morphospace by scaling, rotating, and translating the landmark coordinates [51]. As the resulting Procrustes coordinates still contained an allometric effect, we computed the centroid size (CS) of the facial configurations of landmarks and used this variable [51] to account for the effect of facial size on shape in all subsequent analyses.

To quantify the amount of facial shape variation attributable to sex, age, facial size, and genetic diagnosis we implemented Procrustes ANOVA and MANOVA tests [52,53] using the function ProcD.lm in the Geomorph R package with 1,000 permutations [53,54]. When sex, age or facial size were significant factors influencing facial shape variation, subsequent analyses were adjusted using the residuals derived from the Procrustes MANOVA. Analyses were performed combined and separately for EU and DS samples. When sex differences were detected, further analyses were also performed separately for males and females.

Further Procrustes ANOVA tests were performed to assess whether biomarkers for AD (concentration ratio Aβ1-42/Aβ1-40) and OSA (AHI) explained a significant amount of facial shape variation. Additionally, multivariate linear regressions were performed to further test, statistically and visually, the correlation between facial shape, age, concentration ratio Aβ1-42/Aβ1-40, and AHI.

To assess statistically significant facial shape differences between diagnostic groups, we computed the Procrustes distances between the average shapes of each group and performed permutation tests with 10,000 iterations [50]. Facial comparisons were performed between the following diagnostic groups: a) DS/EU, b) Alzheimer/ No Alzheimer for EU and DS groups, and c) no OSA/mild OSA/severe OSA in the DS group. To visualize the facial shape differences between groups, we conducted Canonical Variate Analysis (CVA), a statistical discriminant method that maximizes the separation among predefined groups [55,56]. Facial morphings associated with the negative and positive extremes of the first two CVs were created with IDAV Landmark Editor version 3.0. Moreover, we created heatmaps using Amira 5.2.1 (v6.3, Visualization Sciences Group, FEI) to identify the facial regions where differences between diagnostic groups were mainly located.

All GM and multivariate statistical analyses were executed using *MorphoJ version 1.07a b* [57] and *Geomorph R-Package version 4.0.5* [54].

## Results

### Sex, age and facial size influence facial shape

Procrustes ANOVA analyses revealed that sex (*Z* = 4.0879, *P* = 0.001), age (*Z* = 7.4097, *P* = 0.001) and facial size (*Z* = 7.1928, *P* = 0.001) were significant factors influencing facial shape in our sample (Additional file 4: Table S3). After adjusting for these factors, the Procrustes MANOVA test also indicated a significant influence of trisomy on facial shape variation (*Z* = 6.9455, *P* = 0.001) (Additional file 5: Table S4). Comparative shape analysis based on Procrustes distances confirmed significant facial differences between EU controls and individuals with DS (*P* < 0.0001), whereas the CVA showed that trisomic individuals exhibited shorter and wider faces, midfacial retrusion with flattened nasal bridges, and oblique palpebral fissures (Fig. 2A), which are the facial features typically associated with DS.

**Fig. 2.**
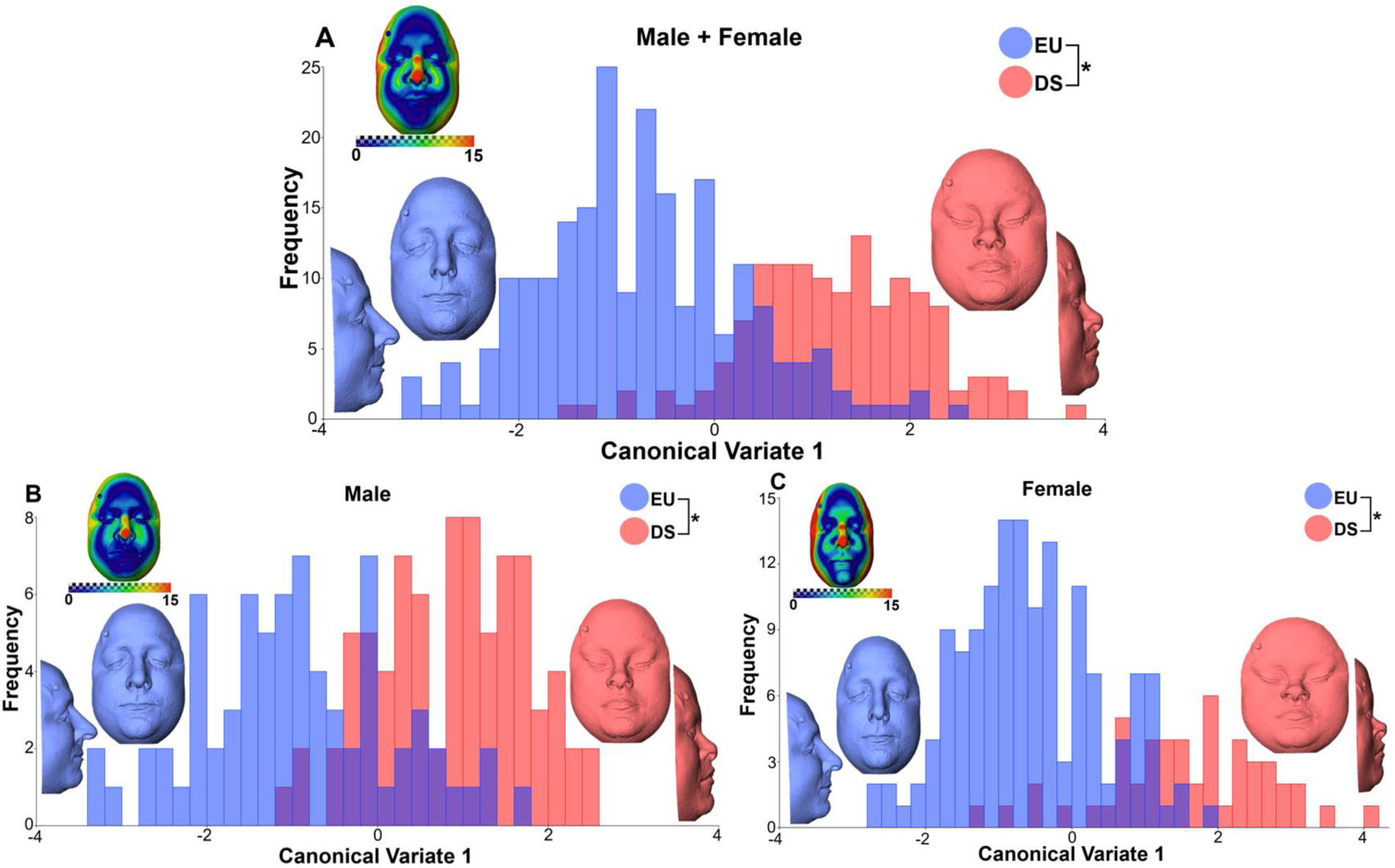
Results of the Canonical Variate Analysis (CVA) between EU controls and individuals with DS. A) Male + Female, B) Male and C) Female groups. The X-axis displaying Canonical Variate1 (CV1) represents 100% of facial shape variation. Facial morphings associated to the negative extreme of CV1 represent frontal and lateral views of the shape changes in EU individuals (blue), whereas facial morphings on the positive extreme represent individuals diagnosed with DS (red). Facial morphings are magnified by a factor of 2. Heatmaps showing facial shape differences between the morphings associated to the positive and negative extreme of CV1. Red areas indicate anatomical regions with largest facial shape differences between EU controls and individuals with DS, whereas blue areas indicate smallest facial shape differences. *Statistically significant difference between groups (*P* <0.0001). The faces in the figure are morphings which do not belong to a real individual and are based on 3D landmark coordinates.

### Sex-dependent effect of trisomy 21 on facial shape

To determine whether trisomy 21 differently affected males and females, we performed a Procrustes MANOVA adjusting only for age and facial size. The results showed that the interaction between genetic diagnosis and sex was statistically significant (*Z* = 3.7046, *P* = 0.002) (Additional file 6: Table S5), suggesting a sex-dependent effect of trisomy 21 on facial shape.

To further explore these sex-specific effects, we performed independent analyses for males and females. Procrustes distances between diagnostic groups confirmed significant facial shape differences between individuals with DS and EU controls in both males (*P* < 0.0001) and females (*P* < 0.0001). The CVAs showed that individuals diagnosed with DS from both sexes showed the typical DS facial features (Fig. 2B and 2C). However, the heatmaps representing facial shape differences between EU controls and DS individuals showed that DS facial traits were more pronounced in females (Fig. 2C).

### Altered facial ontogenetic trajectories in females with Down syndrome

After evaluating the effect of sex on facial shape, we compared how facial shape changes over adulthood in EU and DS groups. A multivariate linear regression analysis revealed a significant relationship of facial shape with age (%Predicted = 7.11%, *P* = <0.0001), and showed the effects of ageing in both groups. Since the slope of the regressions were not parallel, the analysis suggested differences in the ontogenetic trajectories of EU controls and individuals diagnosed with DS (Fig. 3A). A Procrustes MANOVA confirmed a significant interaction between genetic diagnosis and age after adjusting for sex and facial size (*Z* = 1.5826, *P* = 0.005) (Additional file 7: Table S6A), demonstrating that facial adult ontogeny is altered in DS. Individuals with DS presented a steeper slope of the ontogenetic trajectory, showing more pronounced facial shape changes during ageing in comparison to EU controls (Fig. 3A).

**Fig. 3.**
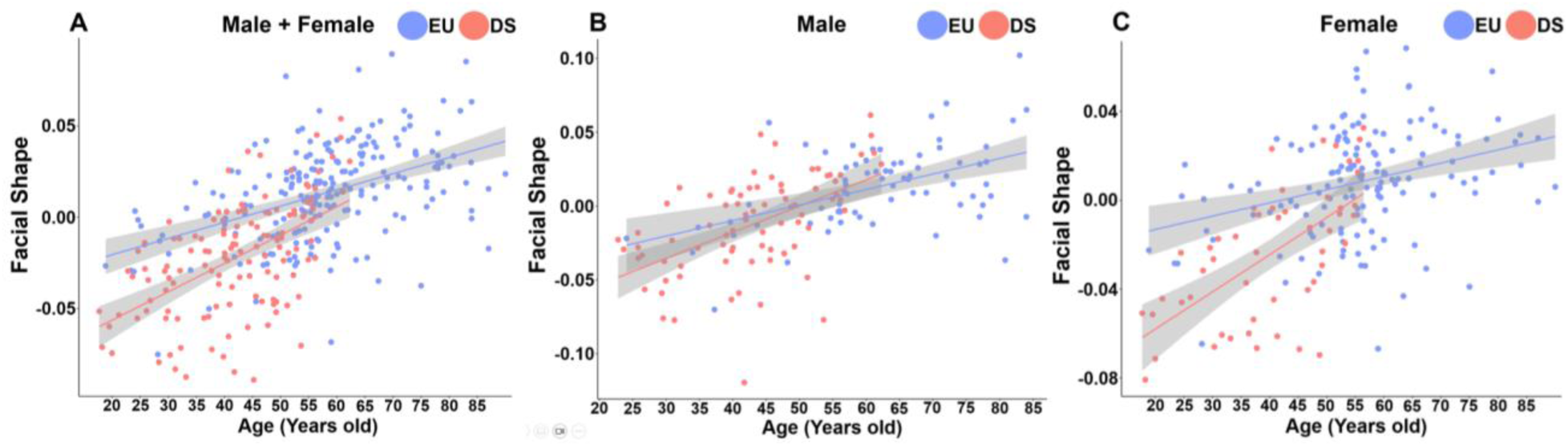
Ontogenetic trajectories of EU and DS populations assessed by multivariate regression of facial shape on age. A) Male + Female, B) Male and C) Female groups.

To further explore sex-specific facial shape changes in the ontogenetic trajectories between EU controls and individuals with DS, we repeated the multivariate regressions and Procrustes MANOVA tests separately for males and females, adjusting only for facial size. In the male sample, the multivariate regression results showed a significant correlation between facial shape and age among EU and DS individuals (%Predicted = 8.79%, *P* = <0.0001) (Fig. 3B). However, the Procrustes MANOVA showed that the interaction between diagnosis and age was not significant (*Z* = -0.3126, *P* = 0.605) (Additional file 7: Table S6B, Fig. 3B), indicating no differences in the ontogenetic trajectories between the EU and DS groups in males. On the contrary, in the female sample, we detected both a statistical significance in the correlation between facial shape and age (%Predicted = 6.98%, *P* = <0.0001) (Fig. 3C), and in the interaction between diagnosis and age (*Z* = 1.8775, *P* = 0.033) (Additional file 7: Table S6C, Fig. 3C). These results indicated that the altered ontogenetic trajectory in the DS population (Fig. 3A) was only due to female individuals (Fig. 3C), who showed more pronounced facial ageing effects.

### Significant correlations of facial shape with AD and OSA

Considering the previous results, we accounted for the effect of sex, age and facial size in the subsequent analyses assessing the potential of facial shape as a diagnostic biomarker for AD and OSA in the DS and EU populations.

Regarding AD, we first investigated the association between facial shape variation with molecular biomarkers of dementia. A multivariate regression analysis indicated that the concentration ratio of Aβ1-42/Aβ1-40 in CSF was significantly correlated with facial shape after adjusting for sex, age and facial size when EU controls and individuals with DS were pooled together (%Predicted = 4.4%, *P* = 0.001) (Additional file 8: Table S7, Fig. 4). However, when the association was evaluated separately for each group, the multivariate regressions did not show significant correlations neither for EU controls (%Predicted = 1.8%, *P* = 0.369) nor for individuals with DS (%Predicted = 2.1%, *P* = 0.689).

**Fig. 4.**
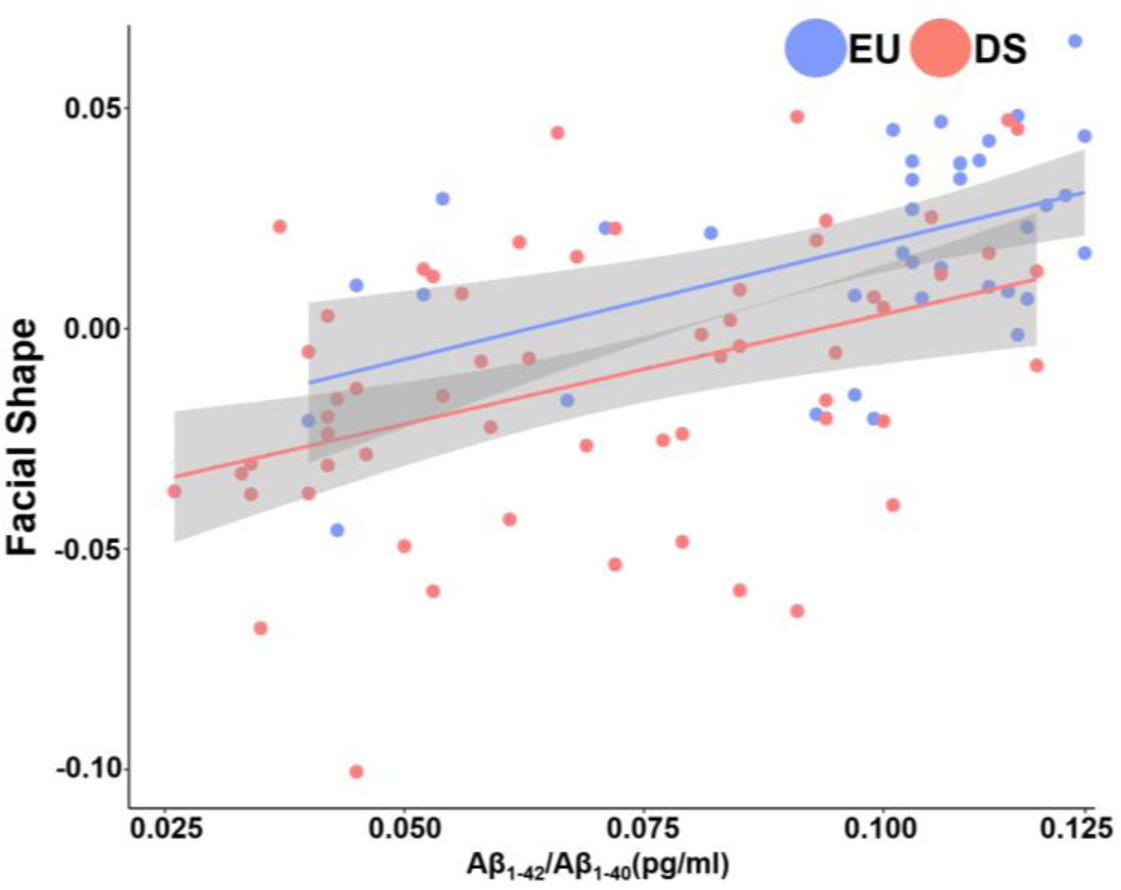
Multivariate regression of facial shape on concentration ratio Aβ1-42/Aβ1-40, an AD diagnostic biomarker.

Second, we explored the facial morphological variation associated with the diagnosis of AD in each group, and separately for males and females. In the DS population, facial shape differences between individuals with and without AD diagnosis were significant before adjusting for age, sex and facial size (*Z* = 2.3385, *P* = 0.008). However, once adjusted for those variables, statistical significance was not achieved (*Z* = -0.30077, *P* = 0.611) (Fig. 5A). Moreover, when we performed the analyses separately for each sex, we did not detect any significant facial shape differences between individuals with and without AD in males (*Z* = 0.21119, *P* = 0.411) (Fig. 5B), nor in females (*Z* = 0.072242, *P* = 0.468) (Fig. 5C).

**Fig. 5.**
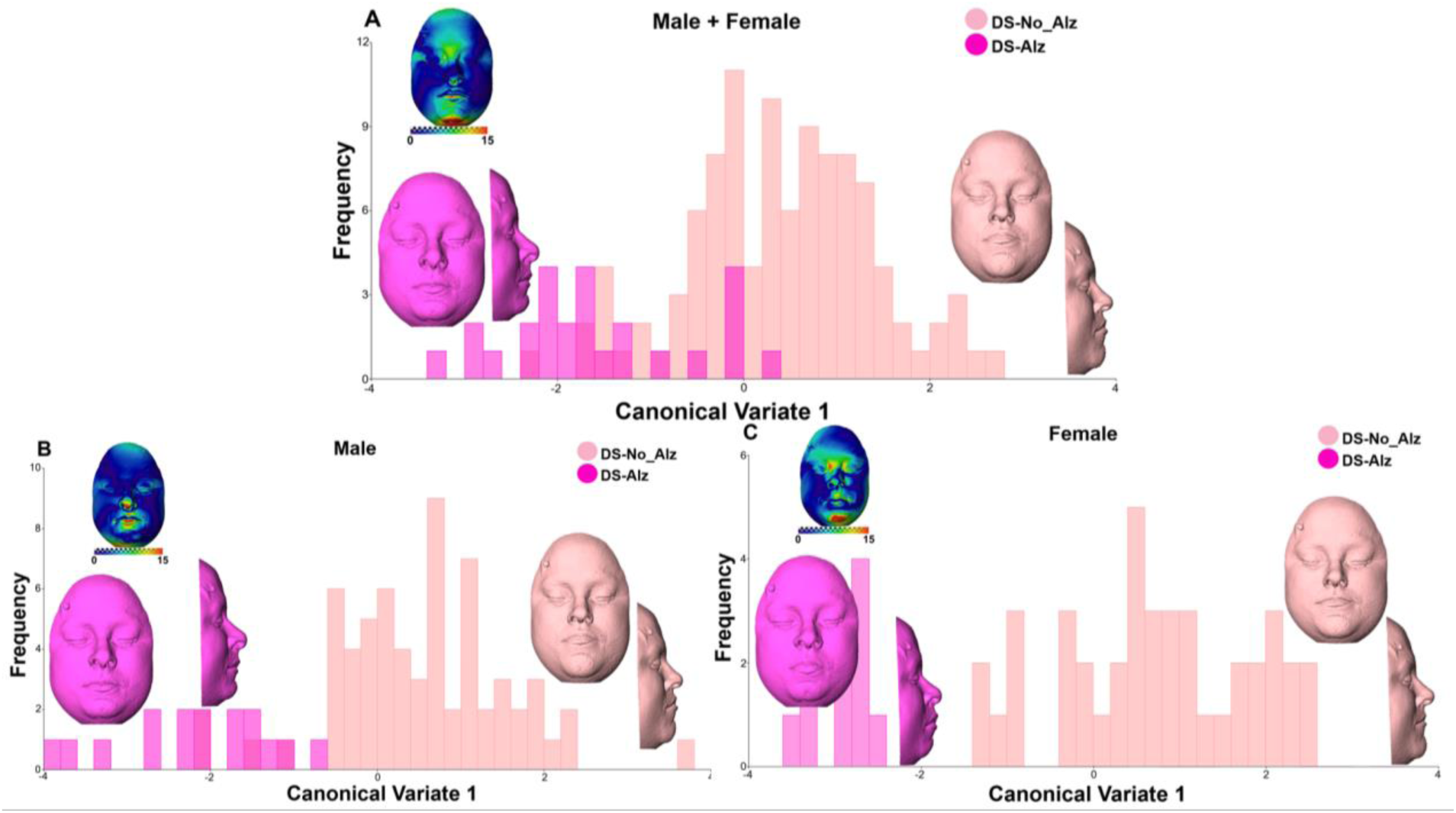
Results of the CVA between DS individuals with and without AD dementia. A) Males + Females, B) Males and C) Females. The X-axis displaying Canonical Variate1 (CV1) represents 100% of facial shape variation. Facial morphings associated to the negative extreme of CV1 represent frontal and lateral views of the shape changes in DS individuals with AD dementia (purple), whereas facial morphings on the positive extreme represent DS individuals without AD dementia (light pink). Facial morphings are magnified by a factor of 2. No statistically significant differences were detected between groups. The faces in the figure are morphings which do not belong to a real individual and are based on 3D landmark coordinates.

In the EU population, significant shape differences were detected between individuals with and without AD diagnosis before (*Z* = 4.4324, *P* = 0.001) and after (*Z* = 3.5777, *P* = 0.001) (Fig. 6A) adjusting for age, sex, and facial size. We found that the main facial morphological differences between EU controls with and without AD were independent from age, with facial changes located in the facial midline, specifically on the forehead and chin (Fig. 6A). Moreover, we identified significant facial differences between individuals with and without AD dementia in both males (*Z* = 2.2095, *P* = 0.015) (Fig. 6B), and females (*Z* = 2.4276, *P* = 0.009) (Fig. 6C). In males, the main facial differences were detected in the facial midline (Fig. 6B), whereas in females the differences were localized on the forehead and the zygomatic prominences (Fig. 6C).

**Fig. 6.**
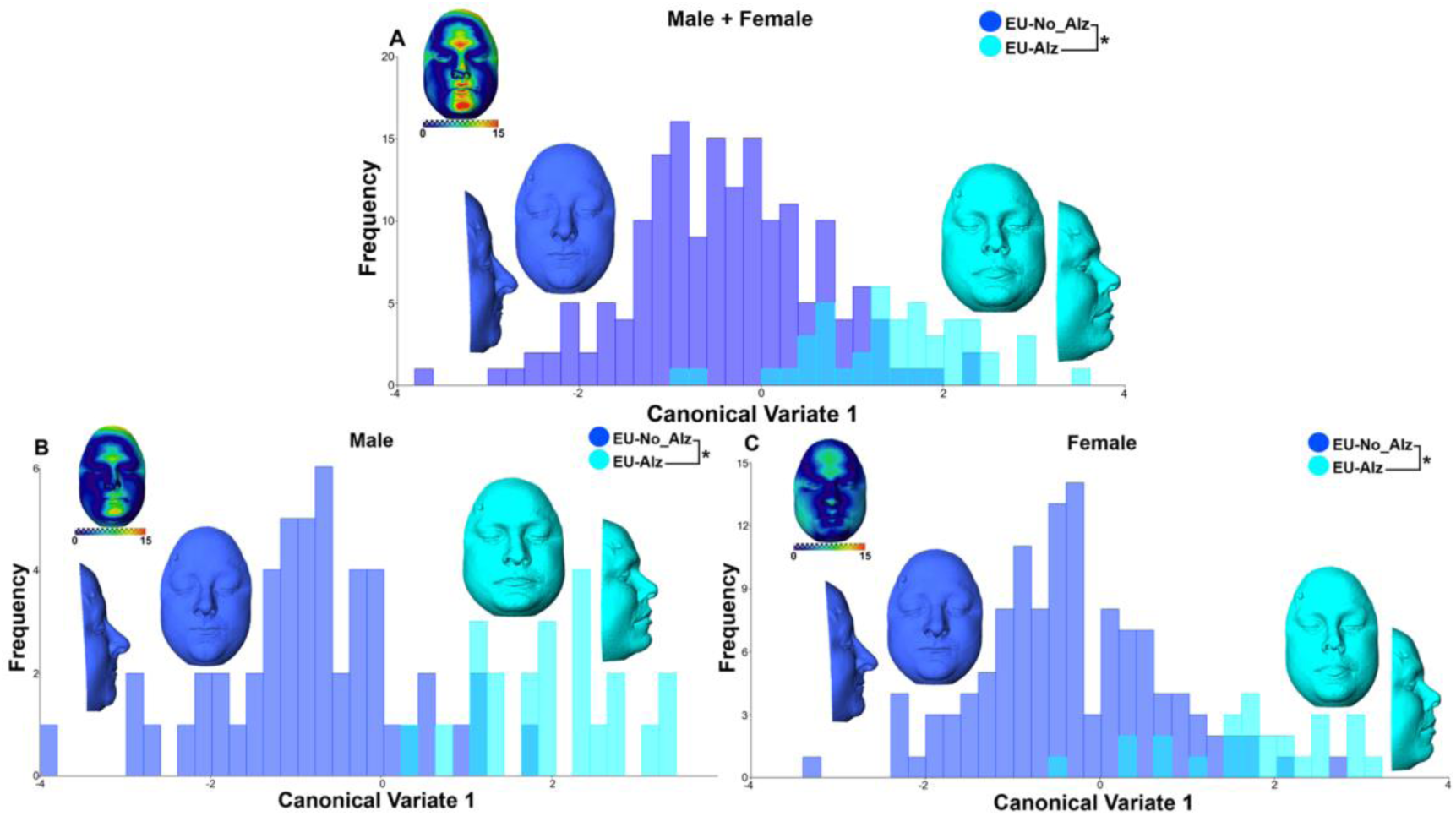
Results of the CVA between EU controls with and without AD dementia. A) Males + Females, B) Males and C) Females. The X-axis displaying Canonical Variate1 (CV1) represents 100% of facial shape variation. Facial morphings associated to the negative extreme of CV1 represent frontal and lateral views of the shape changes in the EU controls without AD dementia (dark blue), whereas facial morphings on the positive extreme represent EU controls diagnosed with AD dementia (light blue). Facial morphings are magnified by a factor of 2. *Statistically significant differences between groups (*P* <0.0001). The faces in the figure are morphings which do not belong to a real individual and are based on 3D landmark coordinates.

Regarding OSA, we explored the association between facial shape variation and a clinical score of sleep apnea in a subsample of individuals. The multivariate regression of facial shape variation on AHI after adjusting for age, sex, and facial size demonstrated a significant correlation when the EU and DS populations were pooled together (%Predicted = 3.4%, *Z* = 1.8139, *P* = 0.039) (Fig. 7). When the association was tested separately for each group, the multivariate regressions indicated no significant correlation for EU controls (%Predicted = 2.1%, *P* = 0.902), but a significant correlation for individuals with DS (%Predicted = 2.9%, *P* = 0.009). A Procrustes ANOVA confirmed that AHI accounted for a small but significant percentage of facial shape variation in the DS group (*Z* = 2.1487, *P* = 0.017) (Additional file 9: Table S8).

**Fig. 7.**
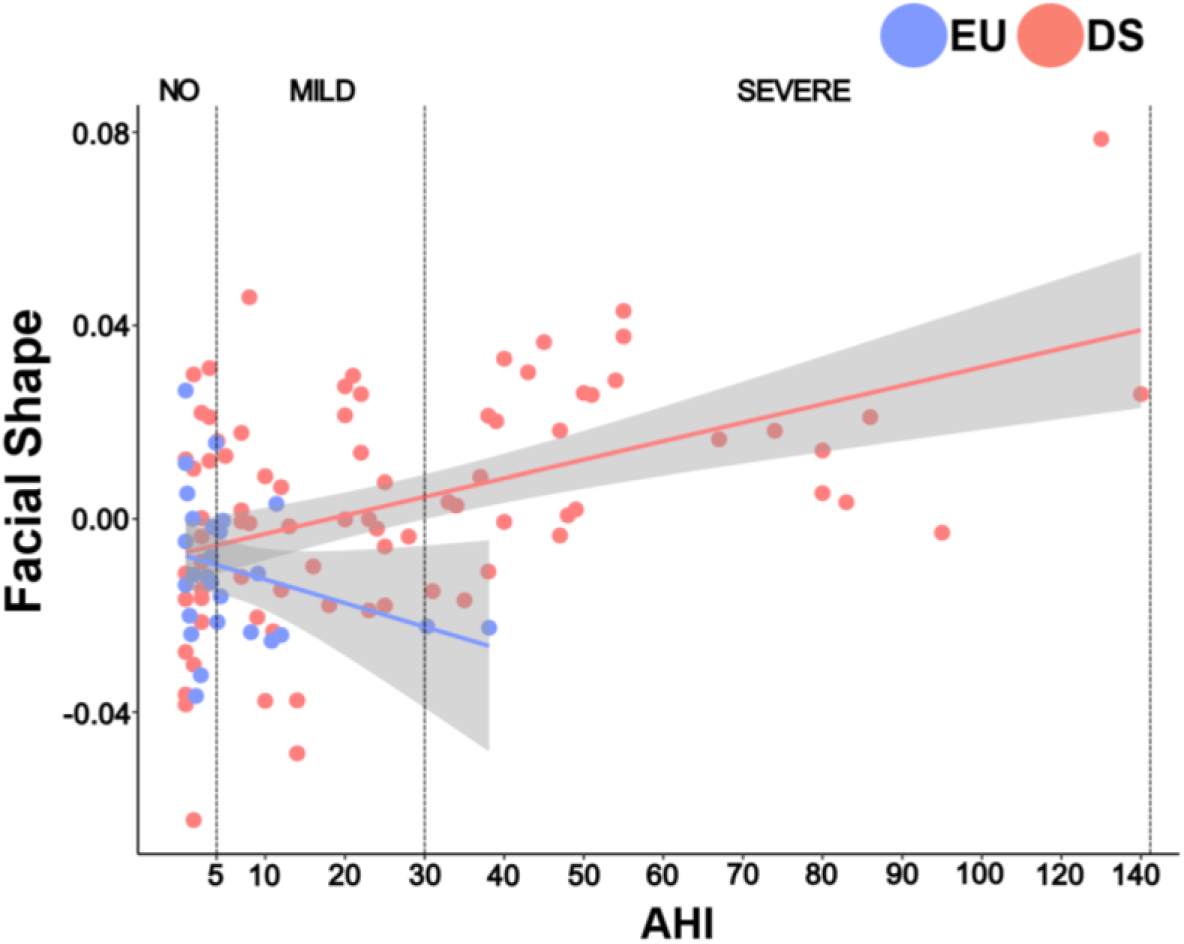
Multivariate regression of facial shape on AHI, an OSA diagnostic biomarker.

Finally, when we assessed whether there were significant facial shape differences between individuals diagnosed with DS and different levels of OSA severity, the Procrustes distances between groups revealed that individuals with DS and severe OSA presented a significantly different facial morphology than individuals with DS and no OSA (*P* = 0.005). The CVA showed that, in comparison to individuals with no signs of OSA, individuals with DS and severe OSA were characterized by narrower faces and subtle mandibular retraction (Fig. 8). Individuals with mild OSA fell between the individuals with no signs of OSA and individuals with severe OSA, showing a continuum spectrum of facial shape variation associated to OSA and suggesting a relationship between facial morphology and OSA worsening.

**Fig. 8.**
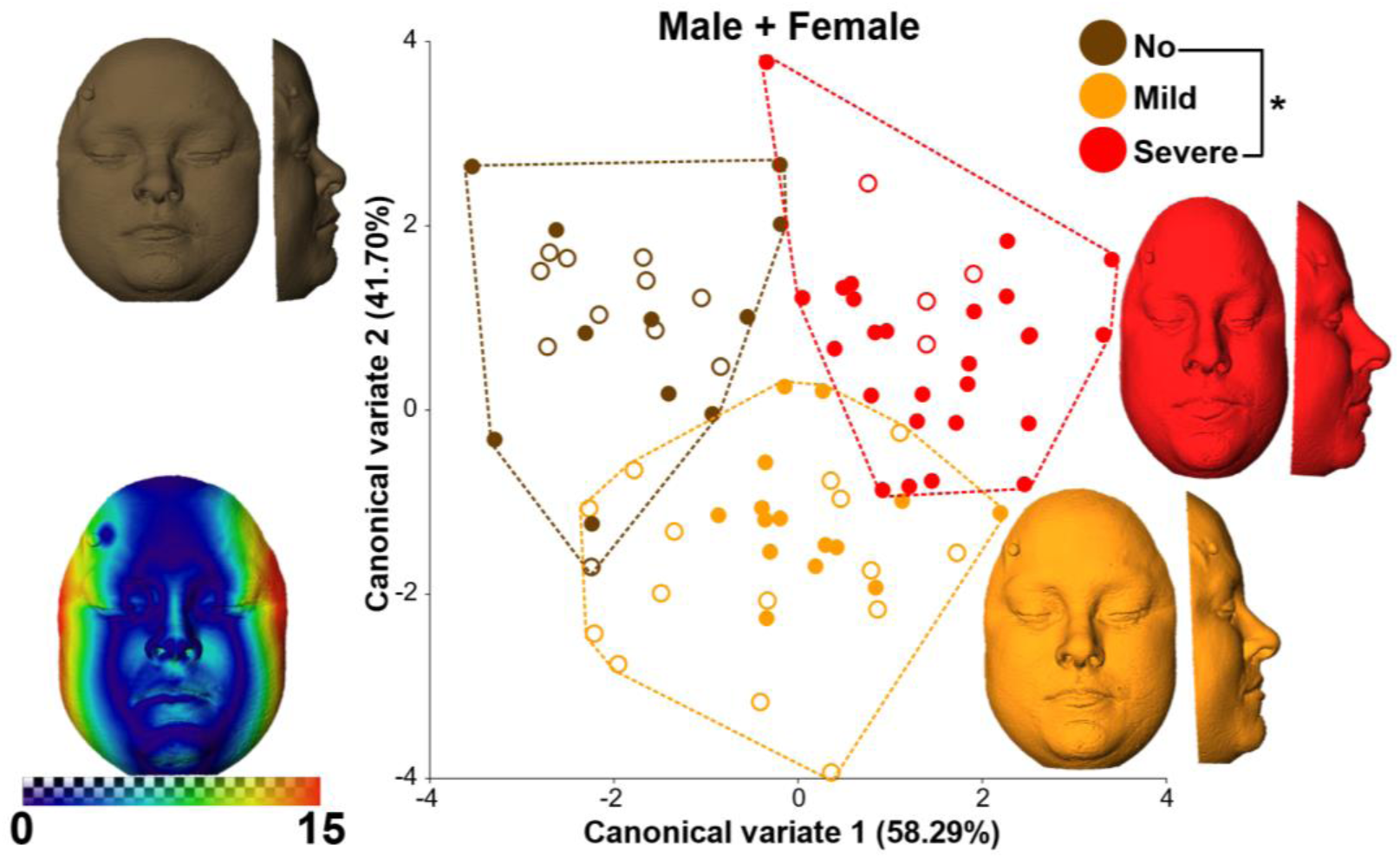
Results of the CVA assessing facial differences between individuals with DS and different OSA levels. Facial morphings (magnified by a factor of 2) corresponding to the consensus morphology/average shape of each group. *Statistically significant differences between groups (P <0.0001). The faces in the figure are morphings which do not belong to a real individual and are based on 3D landmark coordinates.

## Discussion

Our study is the first to perform a comprehensive and quantitative 3D analysis of facial shape variation in Down syndrome (DS) and euploid control (EU) populations to assess the diagnostic potential of facial biomarkers for Alzheimer’s disease (AD) and Obstructive Sleep Apnea (OSA). Our results demonstrated that age, sex, and facial size are significant factors on facial shape variation, and that these factors should be accounted to optimize the diagnostic potential and accuracy of facial biomarkers. Furthermore, we demonstrated that facial shape showed diagnostic and sex-specific patterns. First, we identified more severe facial traits in females with DS. Second, we detected significant facial patterns associated to AD in EU, and to OSA in DS populations. Overall, our results support the use of facial shape as indirect biomarker for neurodegenerative and respiratory disorders in future studies and clinical applications.

Our findings showed that after adjusting for the significant effect of age, sex, and facial size, individuals diagnosed with DS displayed a significantly different facial morphology as compared to EU controls. The facial traits associated with DS in our study coincided with the typical DS facial features described in previous studies [10,11,58], including shorter and wider faces, midfacial retrusion, flattened nasal bridges and oblique palpebral fissures. However, our study provided original evidence that trisomy 21 differentially affects the facial morphology of males and females diagnosed with DS, with females showing more pronounced facial shape differences. Other studies have reported sex-related differences in individuals DS, such as an earlier decline in bone mineral density (BMD) in males [59,60] and a slower rate of BMD acquisition in females [61]. Additionally, females with DS tend to have higher fat levels and lower lean mass, which carries significant health implications [62]. Given that facial anatomy is composed of both bone and soft tissue (fat and lean mass), these previous findings [59,60,61,62] suggest that the reduced BMD and lean mass in females could contribute to the more pronounced DS-associated facial features observed in this population.

Our results also evidenced facial shape changes over adult ontogeny in both the EU and DS populations. These findings are consistent with previous studies indicating that age had a significant effect in facial shape in the general [63] and the DS populations [64,65]. Specifically, these studies showed that individuals with DS exhibit specific facial patterns that appear early in life but can vary in adolescence or adulthood, becoming either subtler or more pronounced [64,65]. Our results showed that adults with DS retained the characteristic facial features associated with trisomy 21, but only females showed a significant deviation of facial adult ontogeny compared to EU controls, displaying more pronounced facial aging effects. Our results thus support previous research showing that even though there is an early genetic contribution establishing the typical facial morphology of diverse syndromes, postnatal ontogeny plays a key role in facial morphological variation throughout life [66]. Moreover, this result aligns with our previous findings, which indicate a more severe phenotype in females with DS, particularly in early adulthood. We detected significant differences in the slopes of the ontogenetic trajectories in females, showing more severe DS phenotypes during adolescence that vary with aging.

Regarding the association between facial morphology and AD, our study demonstrated that facial shape significantly correlated with Aβ1-42 concentration in CSF, which has been proposed as an accurate diagnostic biomarker for AD [19]. Furthermore, we detected significant facial shape differences between individuals with and without AD in the EU population that were sex-specific and independent of age. These results highlight the need for sex-differential approaches for AD detection using facial biomarkers, and suggest that facial morphology could be used as a non-invasive and complementary biomarker for detecting AD. This supports the findings in Umeda-Kameyama et al. (2021) [31] and Zheng et al. (2023) [32], who demonstrated that 2D [31] and 3D [32] facial morphology accurately assists in the non-invasive detection of AD dementia.

However, the significant facial shape differences between individuals with and without AD that were detected in the DS population were not consistent after adjusting for age, sex and facial size. With the available data, we cannot determine whether this result is due to experimental bias from limited sample sizes in the DS population, or due to biological mechanisms associated with DS and AD. It is likely that the facial dysmorphologies caused by trisomy 21 are so pronounced that may mask the facial shape changes associated with AD. Further studies are needed to confirm this hypothesis.

Concerning the association between facial shape and OSA, our study indicated a significant correlation between facial morphology and AHI, the most commonly used biomarker for diagnosing OSA [7]. Furthermore, when analyzing the facial morphological variation associated with OSA, our results demonstrated that individuals diagnosed with DS and severe OSA displayed a significantly different facial shape compared to individuals with DS and no signs of OSA. The facial morphology of individuals with DS affected by severe OSA was characterized by narrower faces and a slight mandibular retraction, which can lead to restricted upper airways and a minor backward displacement of the oropharynx that may explain the higher prevalence of OSA in the DS population. In contrast to previous studies that found no statistically significant differences associated with OSA using 3D photogrammetric measurement [67] or detected only 2D local significant facial differences in the EU population, or using linear measurements based on cephalometric parameters [33,34,35,36]; our study showed significant differences in the DS population. Our results demonstrate that 3D facial anatomy is a potential non-invasive biomarker for detecting OSA and for preventing further cognitive impairment in the DS population.

## Future directions and conclusions

Understanding the bidirectional link between AD and OSA is crucial for the DS population. Several studies have demonstrated that sleep disturbances, including OSA, can increase the risk of an earlier AD onset [68,69], likely due to poor sleep quality, which impairs neuronal repair and reduces the clearance of proteins such as Aβ [14,15] and tau [16,17], leading to increased deposition rates. Moreover, other studies have shown that sleep-related problems generally increase as AD progresses [18,68,70]. Currently, there is no effective cure or treatment for AD that can mitigate the progress of this neurodegenerative disorder. Moreover, people with DS at an ultrahigh risk for suffering AD due to different genetic and morphological factors and invariably will develop AD in adulthood [71,72,73]. However, sleep-related disturbances, such as OSA, display modifiable risk factors that can be treated, minimizing derived neurodegenerative comorbidities such as AD and thereby improving life quality [7,74].

Our results encourage further research using larger samples in the DS population to detect facial shape differences between individuals with and without AD dementia. Regarding OSA, larger samples could enable the evaluation of facial shape differences by sex and to determine if significant morphological differences can be detected in individuals with mild OSA. Moreover, additional data of EU controls with OSA are needed to extend our facial morphological analyses to population with sleep-related disorders but no genetic condition.

Overall, our study demonstrated that 3D facial morphology can serve as a potential non-invasive diagnostic biomarker for disorders such as AD and OSA. The findings underscore the importance of adjusting diagnostic models for sex, age, and facial size to optimize the accuracy of these facial biomarkers. Additionally, early detection of disorders in EU or DS populations through facial biomarkers could facilitate better clinical management and improve the quality of life of people with these conditions.

## Availability of data and materials

Raw image data cannot be made available due to restrictions imposed by the ethics approval. Landmark data supporting the findings of this study are available from the corresponding authors upon reasonable request.

## Abbreviations

DS: Down syndrome
EU: Euploid control
AD: Alzheimer’s disease
OSA: Obstructive Sleep Apnea
APP: Amyloid Precursor Protein
Aβ: β-amyloid
NFL: Neurofilament light
CSF: Cerebrospinal Fluid
PSG: Polysomnography
AHI: Apnea-Hypopnea Index
MRI: Magnetic Resonance image
ADNI: Alzheimer’s Disease Neuroimaging Initiative
RARC: Resource Allocation Review Committee
MV-CNN: Multi-View Consensus Convolutional Neural Networks
GM: Geometric Morphometrics
GPA: Generalized Procrustes Analysis
CS: Centroid size
CVA: Canonical Variate Analyses
BMD: Bone Mineal Density

## Acknowledgements

The authors would like to thank all study participants, their families, and caregivers from the DABNI and SPIN cohort for their support of, and dedication to, this research. We also would acknowledge the Fundació Catalana Síndrome de Down (https://fcsd.org/) for global support. We also acknowledge the Alzheimer’s Disease Neuroimaging Initiative (ADNI) for the data shared. Data collection and sharing for this project was funded by the Alzheimer’s Disease Neuroimaging Initiative (ADNI) (National Institutes of Health Grant U01 AG024904) and DOD ADNI (Department of Defense award number W81XWH-12-2-0012). ADNI is funded by the National Institute on Aging, the National Institute of Biomedical Imaging and Bioengineering, and through generous contributions from the following: AbbVie, Alzheimer’s Association; Alzheimer’s Drug Discovery Foundation; Araclon Biotech; BioClinica, Inc.; Biogen; Bristol-Myers Squibb Company; CereSpir, Inc.; Cogstate; Eisai Inc.; Elan Pharmaceuticals, Inc.; Eli Lilly and Company; EuroImmun; F. Hoffmann-La Roche Ltd and its affiliated company Genentech, Inc.; Fujirebio; GE Healthcare; IXICO Ltd.; Janssen Alzheimer Immunotherapy Research & Development, LLC.; Johnson & Johnson Pharmaceutical Research & Development LLC.; Lumosity; Lundbeck; Merck & Co., Inc.; Meso Scale Diagnostics, LLC.; NeuroRx Research; Neurotrack Technologies; Novartis Pharmaceuticals Corporation; Pfizer Inc.; Piramal Imaging; Servier; Takeda Pharmaceutical Company; and Transition Therapeutics. The Canadian Institutes of Health Research is providing funds to support ADNI clinical sites in Canada. Private sector contributions are facilitated by the Foundation for the National Institutes of Health (www.fnih.org). The grantee organization is the Northern California Institute for Research and Education, and the study is coordinated by the Alzheimer’s Therapeutic Research Institute at the University of Southern California. ADNI data are disseminated by the Laboratory for Neuro Imaging at the University of Southern California.

## Author information

Not applicable.

## Authors and Affiliations

Departament de Biologia Evolutiva, Ecologia i Ciències Ambientals (BEECA), Facultat de Biologia, Universitat de Barcelona (UB), Spain.

L.M Echeverry-Quiceno, S. Llambrich, N. Martínez-Abadías

HER - Human-Environment Research Group, La Salle - Universitat Ramon Llull, Barcelona, Spain.

Á. Heredia-Lidón, X. Sevillano

Multidisciplinary Sleep Unit - Respiratory Department, Hospital de la Santa Creu i Sant Pau, Biomedical Research Institute Sant Pau, Institut de Recerca Sant Pau -, Universitat Autònoma de Barcelona, Barcelona, Spain.

S. Giménez

Memory Unit, Department of Neurology, Institut de Recerca Sant Pau - Hospital de la Santa Creu i Sant Pau, Universitat Autònoma de Barcelona, Barcelona, Spain. Centro de Investigación Biomédica en Red en Enfermedades Neurodegenerativas (CIBERNED), Madrid, Spain.

M. Rozalem-Aranha, J. Fortea

Neuroradiology Section, Department of Radiology, Hospital de la Santa Creu i Sant Pau, Universitat Autònoma de Barcelona, Barcelona, Spain.

M. Rozalem-Aranha

Columbia University College of Dental Medicine, New York, United States.

P. Inampudi

Univ. Bordeaux, CNRS, Ministère de la Culture, PACEA, UMR5199, Pessac, France.

Y. Heuzé

## Contributions

LME-Q, YH, XS, JF and NM-A conceived the project. SG and JF recruited participants, performed MR scanning, cognitive evaluations, polysomnography tests, and diagnosed cases with AD and OSA within the DS cohort. MR-A for providing us the magnetic resonance images and the clinical data associated. AH-L, XS and LME-Q developed and validated the automatic landmarking method. LME-Q, SL, PI, YH and NM-A analyzed data and interpreted results. LME-Q, SL and NM-A prepared the manuscript and figures. All authors critically reviewed the manuscript.

## Ethics declarations

Ethics approval and consent to participate

All participants or their legally authorized representatives coming from Hospital de la Santa Creu i Sant Pau signed an informed consent before enrollment with the protocols approved by the local ethics committee of Hospital de la Santa Creu i Sant Pau following the standards for medical research in humans recommended by the Declaration of Helsinki. To use the ADNI database, we followed an application protocol that, after evaluation, provided us access to the clinical and imaging data of ADNI participants, who signed consent forms approved by the Resource Allocation Review Committee (RARC).

## Consent for publication

Alzheimer’s Disease Neuroimaging Initiative (ADNI) provided us the consent for publication after following all the data request process and review the manuscript.

Competing interests

Mateus Rozalem-Aranha Funding:

Alzheimeŕs Association Research Fellowship to promote Diversity (AARFD-21-852492) Mateus Rozalem-Aranha COI:

-Payed consultancy to VERANEX

-Board participation in Masima – Soluçoes em Imagens Médicas LTDA

## Additional information

Publisher’s Note Springer Nature remains neutral with regard to jurisdictional claims in published maps and institutional affiliations.

## Supplementary information

Supplementary material: Additional file 1: Figure. S1 Pipeline to create 3D facial models from MRIs. Additional file 2: Table S1. Acquisition parameters for MR at the Hospital del Mar, Hospital Clinic and the ADNI database. Additional file 3: Table S2. Anatomical definition of facial landmarks used in geometric morphometric and multivariate statistical. Additional file 4: Table S3. Procrustes ANOVA results quantifying facial shape variation attributable to sex, age and facial size. Additional file 5: Table S4. Procrustes MANOVA results quantifying facial shape variation attributable to sex, age, facial size and diagnosis. Additional file 6: Table S5. Procrustes MANOVA results quantifying facial shape variation attributable to diagnosis, sex and their interaction. Additional file 7: Table S6. Procrustes MANOVA results quantifying facial shape variation attributable to the interaction of diagnosis and age. Linear model between EU and individuals with DS for: A) Male + Female, B) Male and C) Female. Additional file 8: Table S7. Procrustes ANOVA results quantifying the amount of facial shape variation attributable to Aβ (1- 42) concentration. Additional file 9: Table S8. Procrustes ANOVA results quantifying the amount of facial shape variation attributable to the AHI.

