## Supplementary Material for "Potential of facial biomarkers for Alzheimer’s disease and Obstructive sleep apnea in Down syndrome and general population"

**Supplementary information**

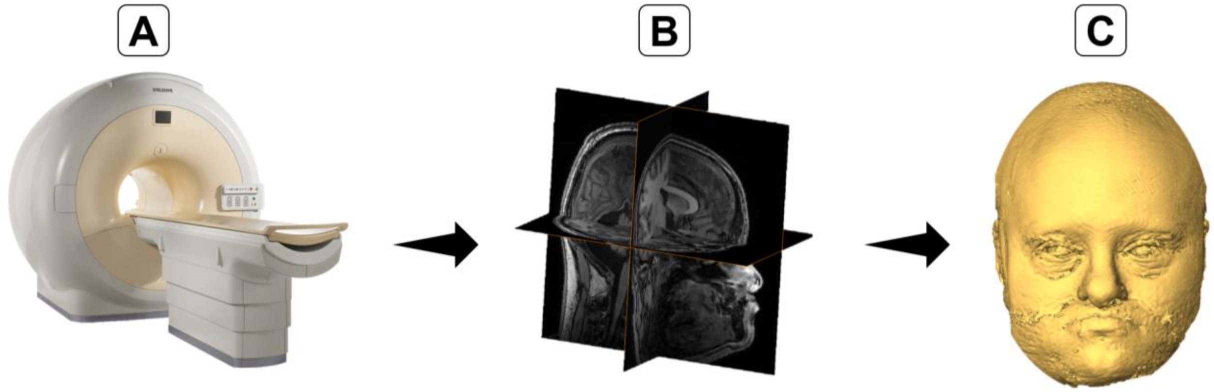

**Additional file 1: Figure. S1** Pipeline to create 3D facial models from MRIs. A) 3T scanner, B) Structural head magnetic resonance and C) 3D facial model.

**Additional file 2: Table S1.** Acquisition parameters for MR at the Hospital del Mar, Hospital Clinic and the ADNI database.

| Database | Matrix size | N° of slices | Voxel size | Echo (TE) | Repetition (TR) | Inversion (TI) | Flip angle |
|---|---|---|---|---|---|---|---|
| Sant Pau Memory Unit | 240×240 | 160 | 1×1×1 $mm^3$ | 3.7ms | 8.1ms | 240ms | 8 |
| ADNI | 240×256 | 250 | 1×1×1 $mm^3$ | 3.0ms | 9.0ms | 230ms | 9 |

**Additional file 3: Table S2.** Anatomical definition of facial landmarks used in geometric morphometric and multivariate statistical.

| Landmark number | Landmark name | Anatomical definition |
|---|---|---|
| 1 | Glabella | Midpoint between the eyebrows on the median plane |
| 2 | Sellion | Deepest point of the nasal root |
| 3 | Pronasale | Most anterior point of the nose tip |
| 4 | Subnasale | Point where the nasal septum meets the philtrum |
| 5 | Labiale superius | Midpoint of the vermelion seam of the upper lip |
| 6 | Labiale inferius | Midpoint of the vermelion seam of the lower lip |
| 7 | Endocanthion R | Point in the internal lateral comissure of the eye (rigth) |
| 8 | Palpebrale inferius R | Most inferior medial point of the lower eyelid (rigth) |
| 9 | Exocanthion R | Point in the external lateral commissure of the eye (rigth) |

| | | |
|---|---|---|
| 10 | Exocanthion L | Point in the internal lateral comissure of the eye (left) |
| 11 | Palpebrale inferius L | Most inferior medial point of the lower eyelid (left) |
| 12 | Exocanthion L | Point in the external lateral commissure of the eye (left) |
| 13 | Alare R | Most lateral point of the nasal wings (rigth) |
| 14 | Subalare R | The facial insertion of the alar base (rigth) |
| 15 | Subalare L | The facial insertion of the alar base (left) |
| 16 | Alare L | Most lateral point of the nasal wings (left) |
| 17 | Chelion R | Point located in the labial commissure (rigth) |
| 18 | Crista philtra R | Crossing of the vermelion (rigth) |
| 19 | Crista philtra L | Crossing of the vermelion (left) |
| 20 | Chelion L | Point located in the labial commissure (left) |
| 21 | Gnathion | Most inferior point of the chin |

**Additional file 4: Table S3.** Procrustes ANOVA results quantifying facial shape variation attributable to sex, age and facial size. The R-squared, the F and Z values, and their statistical significances, are provided. *P-values* were obtained by permutations tests after 10,000 iterations.

| | **EU+DS** | | | | | | |
|---|---|---|---|---|---|---|---|
| | **Df** | **SS** | **MS** | **Rsq** | **F** | **Z** | **Pr(>F)** |
| **Sex** | 1 | 0.04696 | 0.046959 | 0.02079 | 7.3246 | 4.0879 | 0.001 |
| **Residuals** | 345 | 2.21184 | 0.006411 | 0.97921 | | | |
| **Total** | 346 | 2.2588 | | | | | |
| **Age** | 1 | 0.17205 | 0.172051 | 0.07617 | 28.445 | 7.4097 | 0.001 |
| **Residuals** | 345 | 2.08674 | 0.006049 | 0.92383 | | | |
| **Total** | 346 | 2.2588 | | | | | |
| **Facial size** | 1 | 0.13691 | 0.13691 | 0.06061 | 22.26 | 7.1928 | 0.001 |
| **Residuals** | 345 | 2.12189 | 0.00615 | 0.93939 | | | |
| **Total** | 346 | 2.2588 | | | | | |

**Additional file 5: Table S4**. Procrustes MANOVA results quantifying facial shape variation attributable to sex, age, facial size and diagnosis. The R-squared, the F and Z values, and their statistical significances, are provided. *P-values* were obtained by permutations tests after 10,000 iterations.

| | EU+DS | | | | | | |
|---|---|---|---|---|---|---|---|
| | **Df** | **SS** | **MS** | **Rsq** | **F** | **Z** | **Pr(>F)** |
| **Sex** | 1 | 0.05347 | 0.053475 | 0.02367 | 9.989 | 5.2972 | 0.001 |
| **Age** | 1 | 0.06859 | 0.068585 | 0.03036 | 12.812 | 7.2513 | 0.001 |
| **Facial size** | 1 | 0.09695 | 0.096947 | 0.04292 | 18.11 | 6.3908 | 0.001 |
| **Diagnosis** | 1 | 0.09536 | 0.095357 | 0.04222 | 17.812 | 6.9455 | 0.001 |
| **Residuals** | 342 | 1.83086 | 0.005353 | 0.81055 | | | |
| **Total** | 346 | 2.2588 | | | | | |

**Additional file 6: Table S5**. Procrustes MANOVA results quantifying facial shape variation attributable to diagnosis, sex and their interaction. The R-squared, the F and Z values, and their statistical significances, are provided. *P-values* were obtained by permutations tests after 10,000 iterations.

| | EU+DS | | | | | | |
|---|---|---|---|---|---|---|---|
| | **Df** | **SS** | **MS** | **Rsq** | **F** | **Z** | **Pr(>F)** |
| **Diagnosis** | 1 | 0.05835 | 0.058353 | 0.02959 | 10.8661 | 5.6518 | 0.001 |
| **Sex** | 1 | 0.04612 | 0.046115 | 0.02338 | 8.5873 | 5.0265 | 0.001 |
| **Diag:Sex** | 1 | 0.02627 | 0.026267 | 0.01332 | 4.8913 | 3.7046 | 0.002 |
| **Residuals** | 343 | 1.84197 | 0.00537 | 0.93401 | | | |
| **Total** | 346 | 1.9721 | | | | | |

**Additional file 7: Table S6**. Procrustes MANOVA results quantifying facial shape variation attributable to the interaction of diagnosis and age. Linear model between EU and individuals with DS for: A) Male + Female, B) Male and C) Female. The R-squared, the F and Z values, and their statistical significances, are provided. *P-values* were obtained doing resampling permutations tests after 10,000 iterations.

| **A** | Male + Female | | | | | | |
|---|---|---|---|---|---|---|---|
| | **Df** | **SS** | **MS** | **Rsq** | **F** | **Z** | **Pr(>F)** |
| **Diag:Age** | 1 | 0.0099 | 0.009898 | 0.00502 | 1.8022 | 1.5826 | 0.005 |
| **Residuals** | 343 | 1.88385 | 0.005492 | 0.95525 | | | |
| **Total** | 346 | 1.9721 | | | | | |

| | | | | | | | |
|---|---|---|---|---|---|---|---|
| **B** | **Male** | | | | | | |
| | **Df** | **SS** | **MS** | **Rsq** | **F** | **Z** | **Pr(>F)** |
| **Diag:Age** | 1 | 0.00464 | 0.004641 | 0.00472 | 0.797 | -0.3126 | 0.605 |
| **Residuals** | 148 | 0.86187 | 0.005823 | 0.876 | | | |
| **Total** | 151 | 0.98386 | | | | | |
| **C** | **Female** | | | | | | |
| | **Df** | **SS** | **MS** | **Rsq** | **F** | **Z** | **Pr(>F)** |
| **Diag:Age** | 1 | 0.00988 | 0.009876 | 0.0092 | 1.9963 | 1.8775 | 0.033 |
| **Residuals** | 191 | 0.94493 | 0.004947 | 0.88001 | | | |
| **Total** | 194 | 1.07378 | | | | | |

**Additional file 8: Table S7**. Procrustes ANOVA results quantifying the amount of facial shape variation attributable to $A\beta_{1\text{-}42}/A\beta_{1\text{-}40}$ concentration. The R-squared, the F and Z values, and their statistical significances, are provided. *P-values* were obtained by permutations tests after 10,000 iterations.

| | **EU+DS** | | | | | | |
|---|---|---|---|---|---|---|---|
| | **Df** | **SS** | **MS** | **Rsq** | **F** | **Z** | **Pr(>F)** |
| **$A\beta_{1\text{-}42}/A\beta_{1\text{-}40}$** | 1 | 0.0227 | 0.0227026 | 0.04372 | 4.2976 | 3.4105 | 0.001 |
| **Residuals** | 94 | 0.49656 | 0.0052826 | 0.95628 | | | |
| **Total** | 95 | 0.51927 | | | | | |

**Additional file 9: Table S8**. Procrustes ANOVA results quantifying the amount of facial shape variation attributable to the AHI. The R-squared, the F and Z values, and their statistical significances, are provided. *P-values* were obtained by permutations tests after 10,000 iterations.

| | **DS** | | | | | | |
|---|---|---|---|---|---|---|---|
| | **Df** | **SS** | **MS** | **Rsq** | **F** | **Z** | **Pr(>F)** |
| **AHI** | 1 | 0.01343 | 0.0134325 | 0.02736 | 2.2505 | 2.1487 | 0.017 |
| **Residuals** | 80 | 0.47749 | 0.0059687 | 0.97264 | | | |
| **Total** | 81 | 0.49093 | | | | | |
